# Dynamical model for social distancing in the U.S. during the COVID-19 epidemic

**DOI:** 10.1101/2020.05.18.20105411

**Authors:** Shirish M. Chitanvis

## Abstract

**Background:** Social distancing has led to a “flattening of the curve” in many states across the U.S. This is part of a novel, massive, global social experiment which has served to mitigate the pandemic in the absence of a vaccine or effective anti-viral drugs. Hence it is important to be able to forecast hospitalizations reasonably accurately.

**Methods:** We propose on phenomenological grounds a generalized diffusion equation which incorporates the effect of social distancing to forecast the temporal evolution of the probability of having a given number of hospitalizations. The probability density function is log-normal in the number of hospitalizations, which is useful in describing pandemics where the number of hospitalizations is very high.

**Findings:** We used this insight and data to make forecasts for states using Monte Carlo methods. Back testing validates our approach, which yields good results about a week into the future. States are beginning to reopen at the time of publication and our forecasts indicate possible precursors of increased hospitalizations.

Additionally we studied the reproducibility *Ro* in New York (Italian strain) and California (Wuhan strain). We find that even if there is a difference in the transmission of the two strains, social distancing has been able to control the progression of COVID 19.

**Funding:** None.

## I. INTRODUCTION

One goal while tracking the COVID-19 epidemic is to make forecasts of hospitalizations[1]. This calls for caution[2] as the uncertainty in the forecast has to be estimated for a virgin virus. The uncertainty can be reduced by using what has been learned from the history of hospitalizations. It has been reported that the IHME model used such an approach to estimate the maximum hospitalizations in NY state within a factor of two or so.

A straightforward way to make a forecast is to use extrapolation of previous data. One then needs a model of some ilk in order to quantify the uncertainty in the forecast. Uncertainty arises because the transmission of the disease is a probabilistic process which depends on on the distance of closest approach, virus load, time of contact, susceptibility of the target etc. Some models use epidemiological knowledge from previous occurrences of a disease to inform their predictions. In the case of COVID-19 which is a novel virus attacking humans, it is not entirely clear that assumptions from other epidemics apply[3]. Indeed, it would be desirable to have a model for the evolution of hospitalizations which depends only on the history of the current epidemic.

### Added value of this study

We developed a Fokker-Planck/diffusion equation model for the estimation of uncertainty as we extrapolate in time.

We will show via back-testing that our approach yields useful results about a week into the future. We show formally that our approach gives results with narrower uncertainty bands that do standard distributions.

## II. METHODS

A chain reaction model describes an exponential growth in the number of infections (or hospitalizations) *I*(*t*) for the virus:

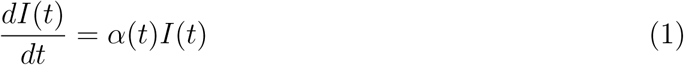

where *α*/ln(2) is the inverse doubling time. *α* can be reduced via social distancing and is related to the reproducibility *Ro:*

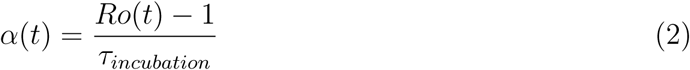

where *τ_incubation_* is the incubation time for a disease. The incubation time for COVID-19 has been quoted to range from a few days to 14 days.

The formal solution to Eqn. 1 is:

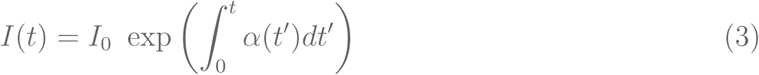

This chain reaction is essentially a probabilistic process, where the chances of transmission between an infected person and a target could depend on the distance of closest approach, virus load or duration of contact, susceptibility of the target etc. The question we would like to address is whether one can obtain a fundamental model to describe the probabilistic progress of the disease.

Towards that end let us focus on the exponent in Eqn. 3. The number of hospitalizations *I* at a given time *t* empirically displays an exponential growth in which the exponent can change in time to reflect the effect of social distancing (see e.g. result for NY state in Fig. 1a, which displays a “square root x” behavior near the beginning of the data):

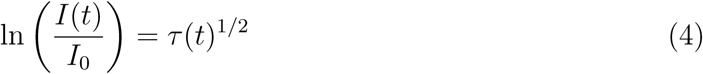

where *τ*(*t*)^1/2^ is a general function which describes the flattening effects of social distancing, or lack thereof.

**FIG. 1.**
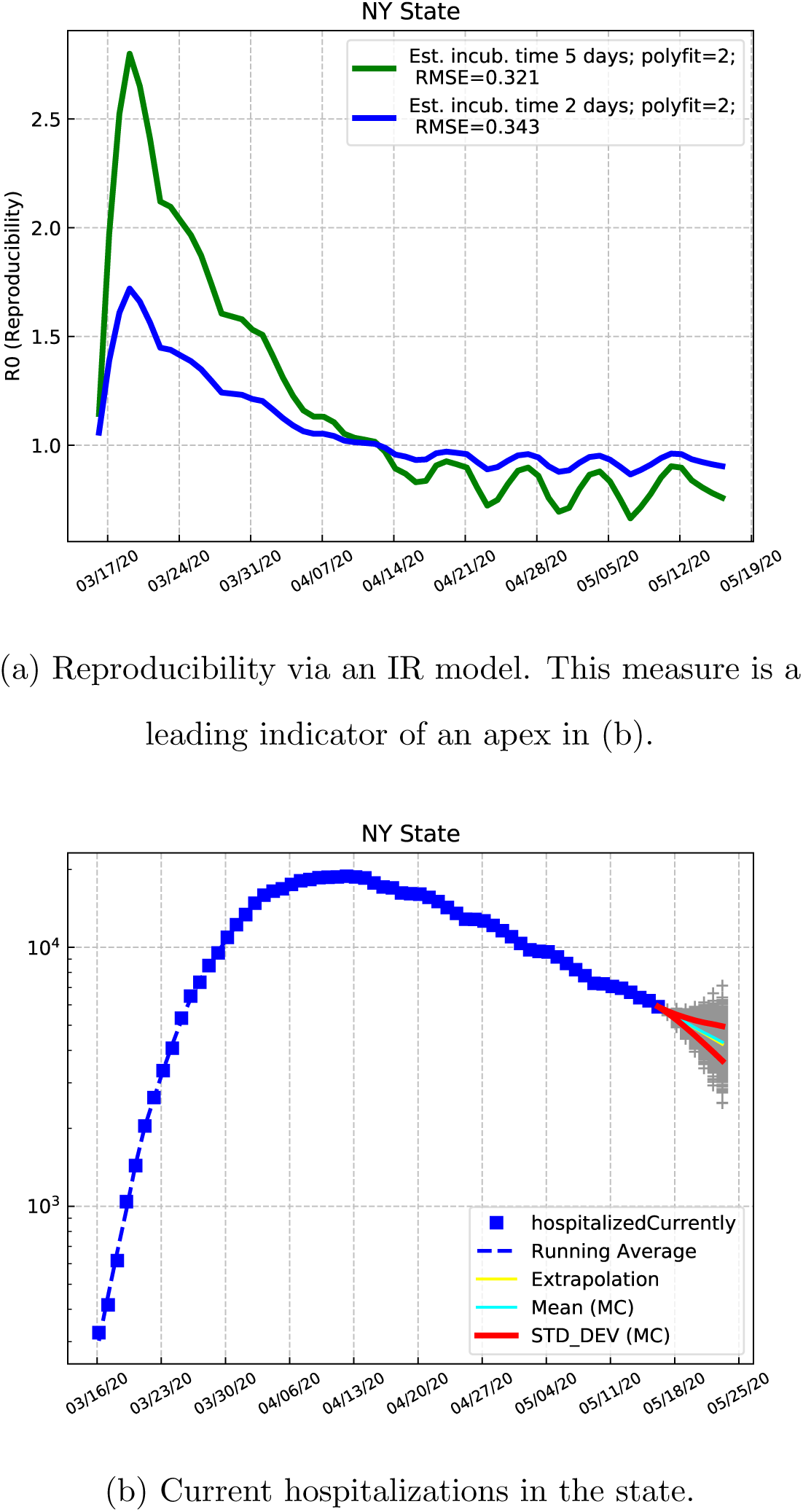

Using data, we can obtain *τ*(*t*)^1/2^ up to some time *t* ≤ *t*_0_. We expect that for a short period of time determined by changes in social distancing behavior for example, we can extrapolate *τ*(*t*)^1/2^ for *t > t*_0_. The disease progresses exponentially, and hence small changes at a given instant in time can have a large effect on later results. This is the reason why we do not expect forecasting to hold beyond a short period.

If *τ*(*t*)^1/2^ ~ *t*^1/2^, the process is analogous to the process of diffusion where the root mean square distance *ξ_rms_* traveled by a particle is proportional to *t*^1/2^. In our case 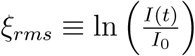. And we know that the probability *P*(*x,t*) of finding a diffusing particle a distance *x* after a time *t* is described by:

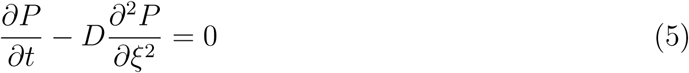

where the diffusion constant is *D*.

Indeed, we found empirically that *τ*(*t*)^1/2^ ~ *t^p^* gave a crude fit to data, with *p <* 0.5 for several states we considered. In our case *ξ* ≡ ln(*I*/*I*_0_), so that the general master equation we seek follows by inspection:

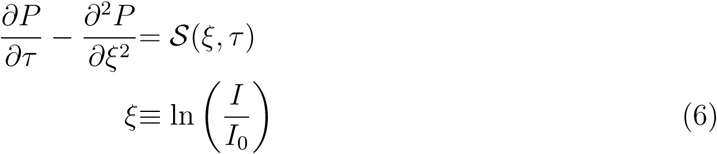

where *τ* ≡ *τ* (*t*), *P* (*ξ*, *τ*) is the probability that *ξ* will have a certain value at a “time” *τ* ≡ *τ*(*t*) [*dimensionless*], while 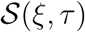 is an arbitrary source function. The diffusion constant is taken to be unity without loss of generality.

If 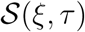 is a Dirac delta function at the *ξ* = *ξ*_0_, *τ* = *τ*_0_, the normalized solution is:

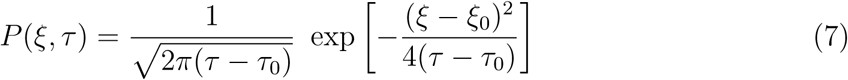

And since *ξ* is logarithmic in *I*, it follows that the number of hospitalizations will be determined by a log-normal distribution. This is especially meaningful since the pandemic shows signs of producing an immense number of infections. We have used Eqn. 7 to make probabilistic predictions of how the number of infections will evolve in time. The variance is given by 2*D*(*τ*(*t*) − *τ*(0)), and the mean is *ξ*_0_. Here the diffusion constant *D* =1 [*dimensionless*].

### A. Data extraction

We used COVID19 state-wide hospitalization data from the Covid tracking project: https://covidtracking.com/api/v1/states/daily.csv to plot and obtain fits to *τ*(*t*)^1/2^.

### B. Data Analysis

A simple IR (Infections (hospitalizations), Recovered cases) model[4], a subset of the SEIR model[5], was used to obtain *Ro* from this data. By dealing directly with a population of hospitalized patients, we are not required to track susceptible and exposed persons. We find from data that the recovery rate *γ* ≈ 0.1[1/*Day*], where *dR/dt* = *γI*. We used Python to perform all the analysis presented in this paper.

The model for extrapolation we have proposed is not unique. There are multiple methods to extrapolate based on previous data. In the case of the novel virus COVID-19 it may be useful to have a diverse set of prediction models to understand the effect of social distancing. The standard probability distribution function used in epidemiology[6, 7] is the Erlang distribution *E_r_*(*k*, *μ*), which is related to the gamma distribution. There are qualitative differences between the standard model and ours. The normal distribution allows the independent specification of the mean and the variance. The Erlang distribution is such that the ratio of the square of the mean (*kμ*) and the variance (*kμ*^2^) is *k*. As such the Erlang distribution will have a large variance if the mean is large, but not necessarily in our log-normal distribution, where the ratio is 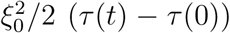. Our model yields variances that are smaller than those obtained from the Erlang distribution. Back testing results for NY state are included in Appendix A which validate our model.

## III. RESULTS

**FIG. 2.**
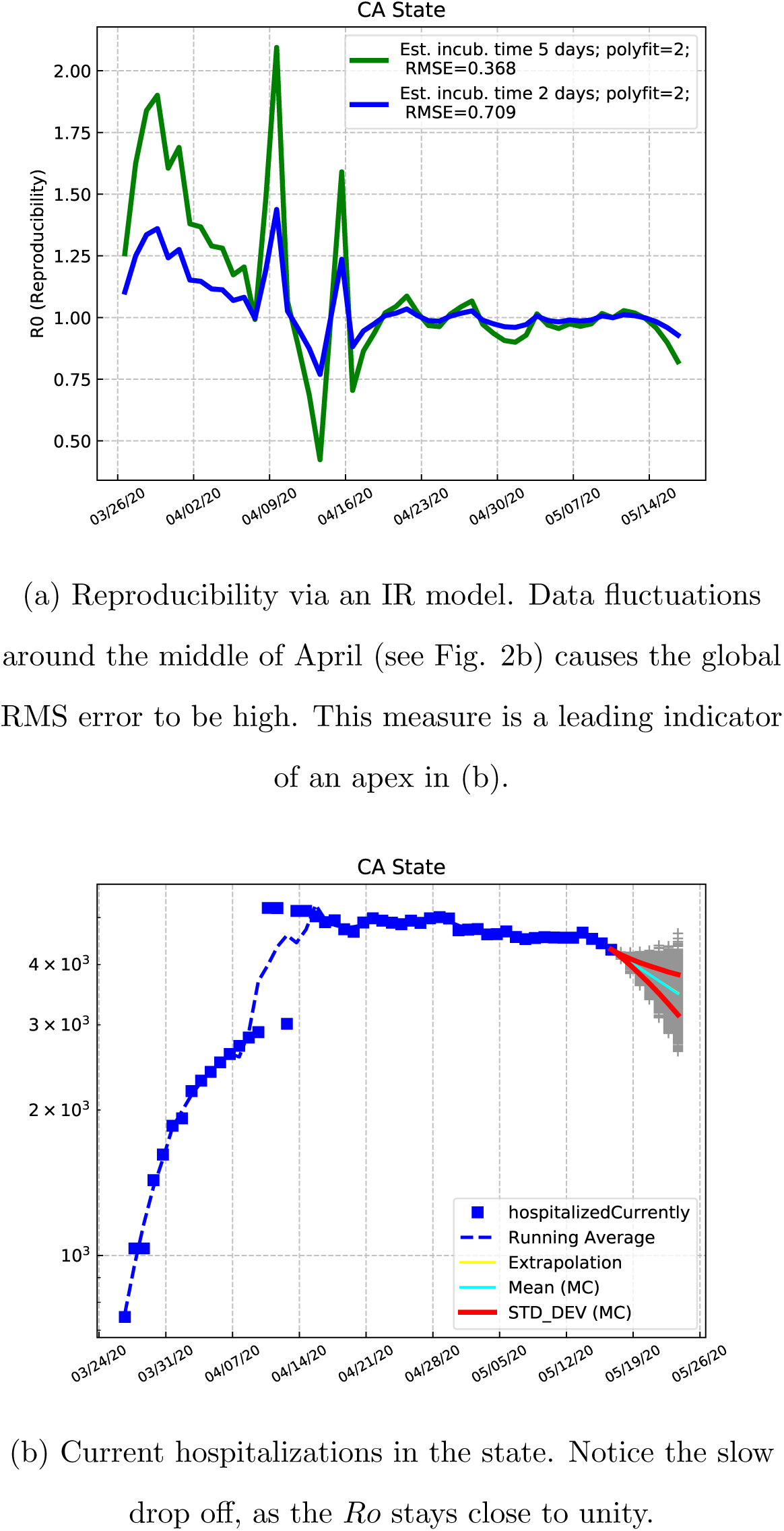

**FIG. 3.**
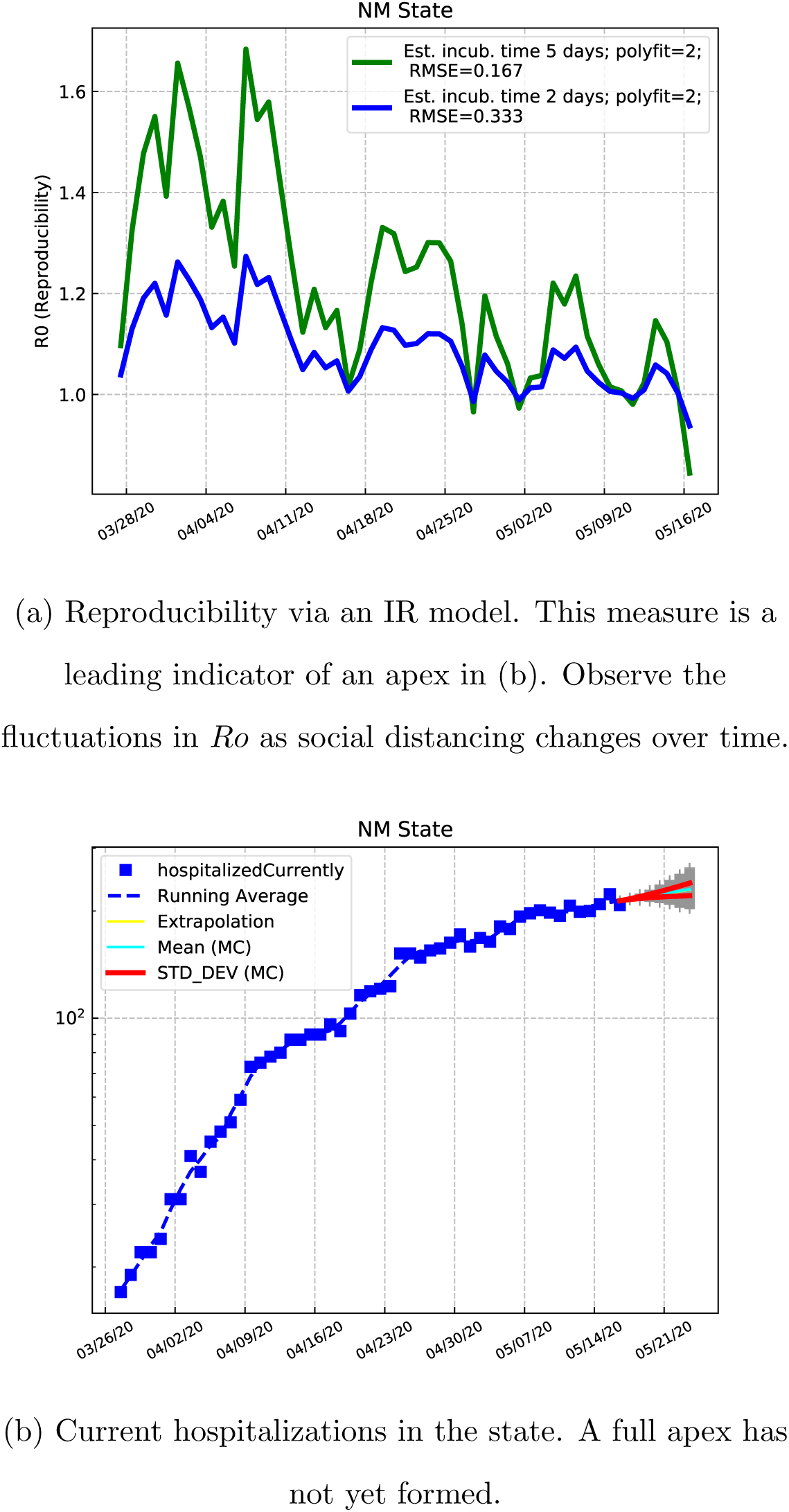

**FIG. 4.**
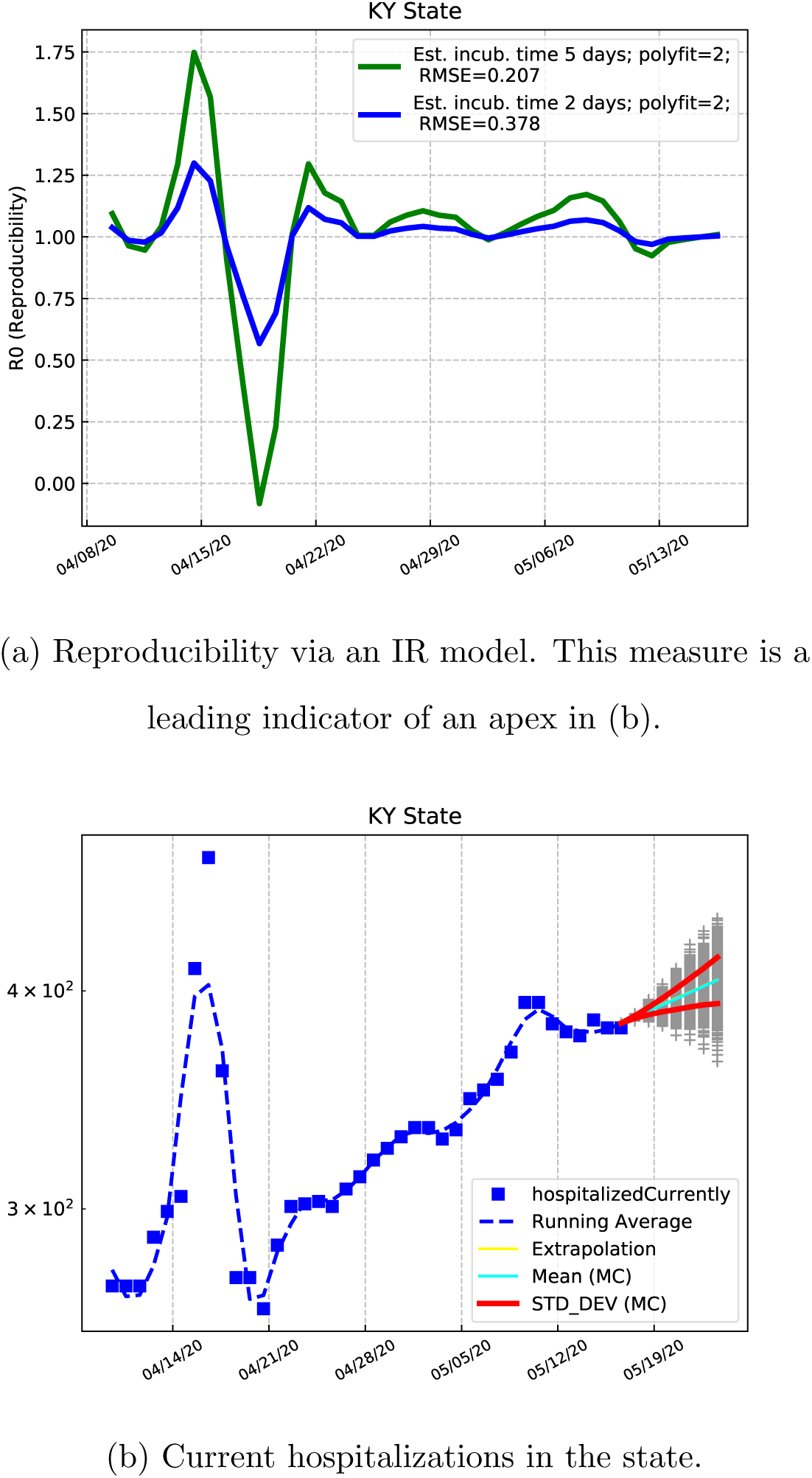

**FIG. 5.**
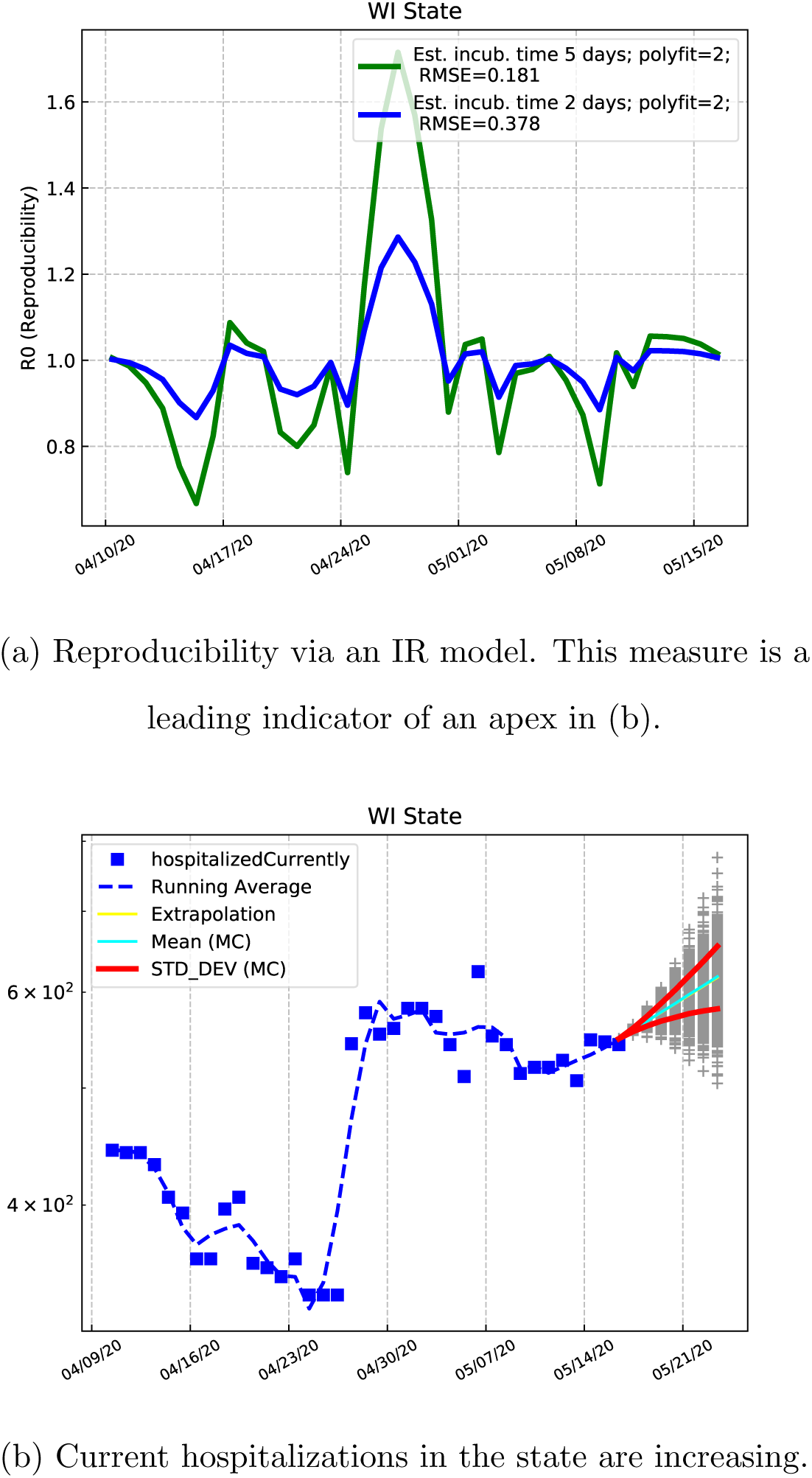

**FIG. 6.**
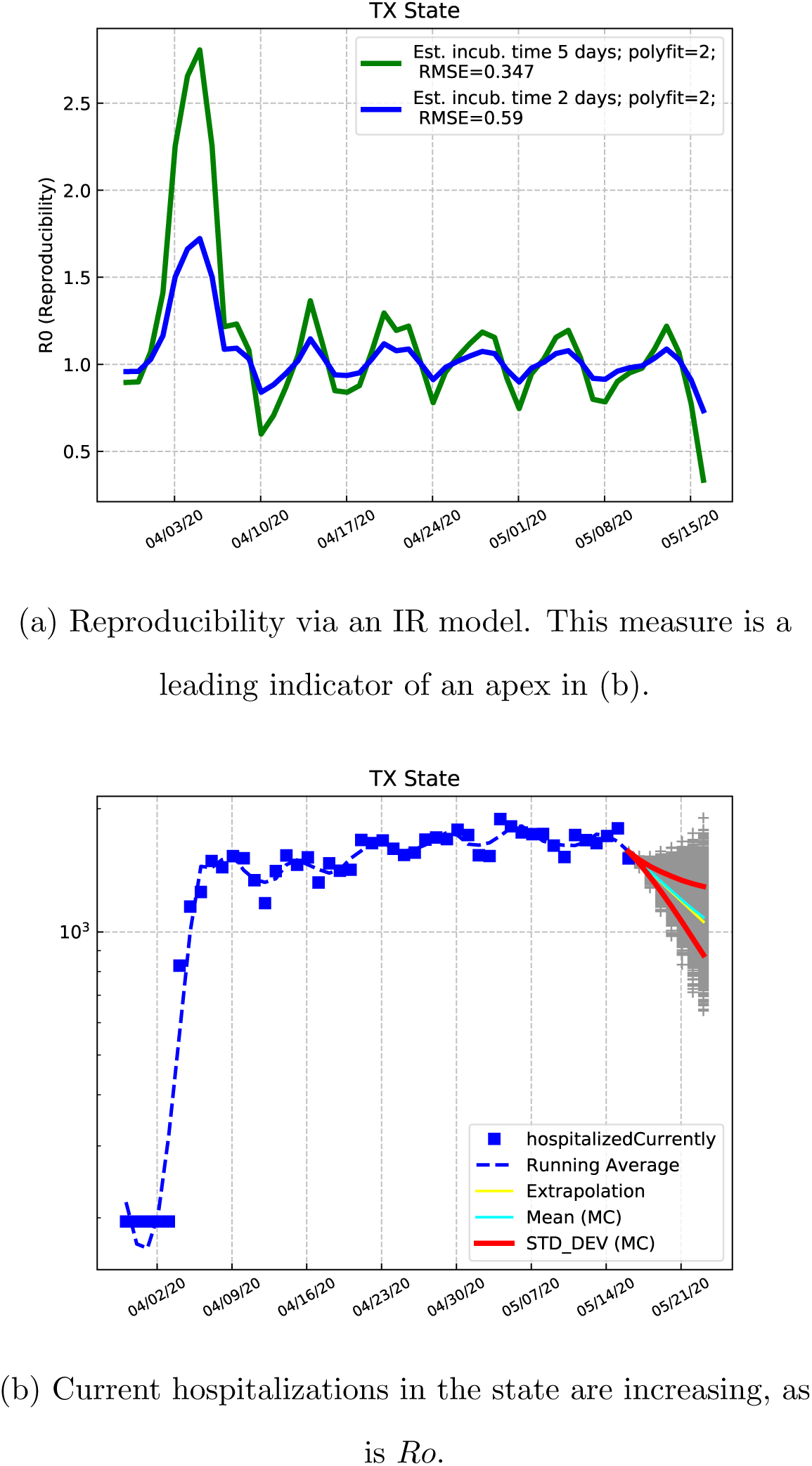

## IV. DISCUSSION

The state of the COVID-19 epidemic is currently fluid. At the time of publication states around the U.S. were beginning to reopen. And you can see possible precursors of resurgence of hospitalIzations in some of them. California continues to reside on a plateau with *Ro* ≈ 1. The reported hospitalizations in Kentucky appear to be relatively low. Observe the intra-week systematic “waves” in hospitalizations for almost every state.

The important point to keep in mind is that the health care system must not be allowed to approach saturation to prevent disastrous situations from developing in a state. This has to be balanced against a difficult decision of what constitutes an “acceptable” casualty rate, before states can be re-opened. While such statements are self-evident, tools like the one we have developed in this paper can be used to inform policy decisions.

A recent preprint[8] suggests that the European strain of COVID 19 may be more transmissible than the Wuhan strain. In light of this we examine the progress of the disease in California (presumably caused by the Wuhan strain), and New York (presumably caused by the Italian strain). Based on a comparison of Figs. 1a and 2a, it may be tempting to say that the higher *Ro* in NY implies a higher transmissibility in that state, compared to CA. But we know that social distancing was imposed in NY later than in CA. So the difference could have arisen for this reason. Furthermore, we cannot distinguish the Ro between the two coasts within the estimated RMS error. In any event, as the curves were flattened, Ro diminishes below one. The implication is that even if there is a difference in the transmission of the two strains, social distancing has been able to control the progression of COVID 19.

## V. CONCLUSION

The main message is that the growth/decay of cases in a pandemic is governed by a lognormal distribution. This distribution changes in time according to a generalized diffusion equation. The log-normal distribution arises from the fact that an epidemic is rather like a chain reaction in a fission bomb.

## Data Availability

Publicly available data on COVID-19 hospitalizations was used from the Covid Tracking Project

https://covidtracking.com/api/v1/states/daily.csv

## VI. ACKNOWLEDGMENTS

I would like to acknowledge discussions with John Scott, and I thank LTC Bahaghidat for encouragement. Last, but not least, I appreciate the references provided to me by LTC Clark.

The work described in this paper was done under the auspices of the DOE while the author was stationed at West Point in the Department of Physics and Nuclear Engineering.

## Appendix A: Back testing to validate our model

We show the results of back testing the time-series analysis for NY, extrapolating seven days. Observe that the 85% confidence bands adequately describe the evolution over one week.

**FIG. 7.**
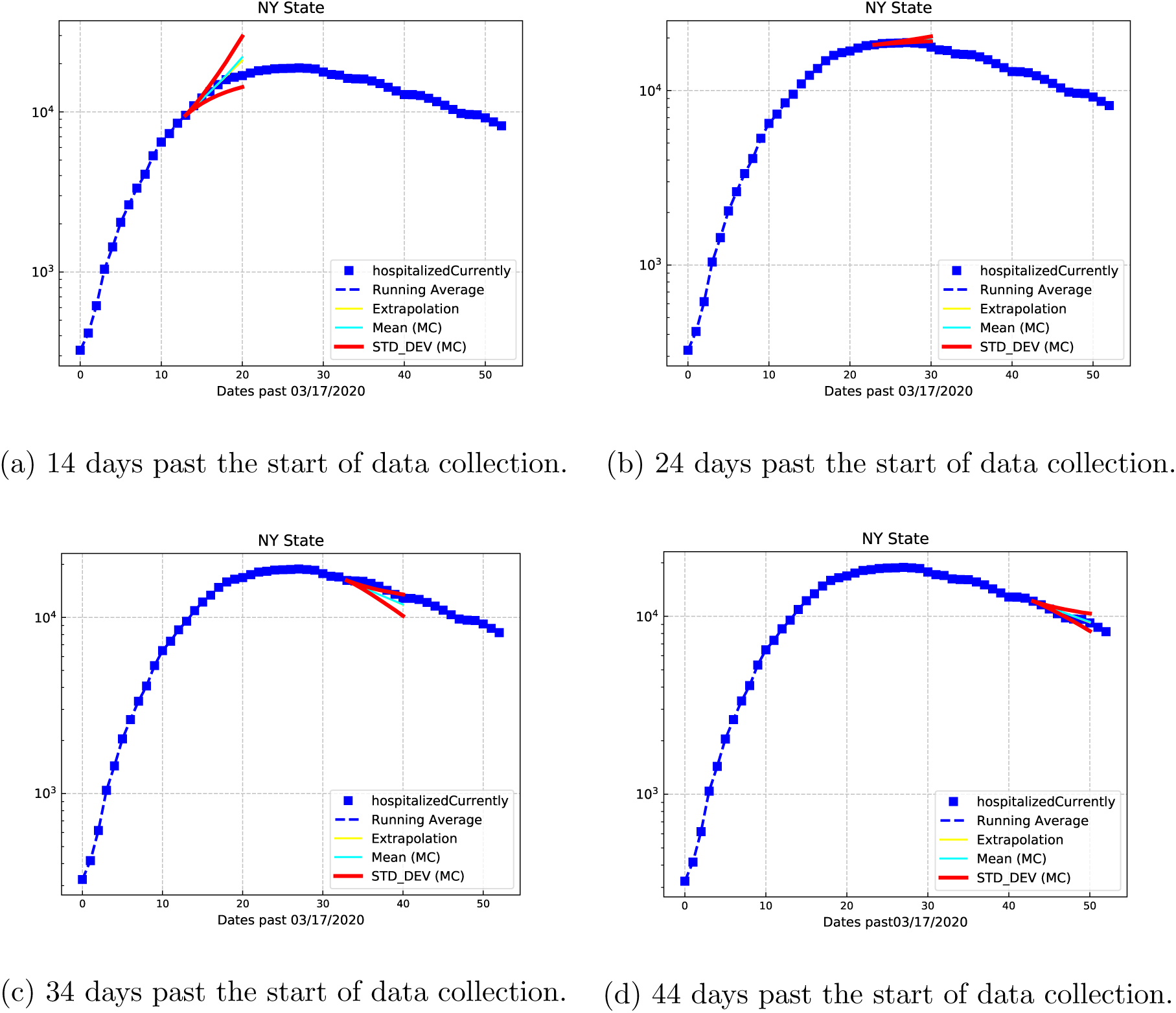

